# SARS-CoV-2 antibody immunity across three continents: the West Africa, West Indies, West London Consortium

**DOI:** 10.1101/2025.06.13.25329588

**Authors:** David Greenwood, Oliver Hague, Eliza Mari Kwesi-Maliepaard, Shanice A Redman, Crick Serology Consortium, the HERITAGE study team, Legacy Investigators, WINDFall Study Team, WWW Consortium Team, Flora Scott, Joshua J Anzinger, Gordon Awandare, David LV Bauer, Yaw Bediako, Edward J Carr, Christine VF Carrington, Adam Kucharski, Peter Quashie, Emma C Wall, Mary Y Wu

## Abstract

**Background:** The experience of the COVID-19 pandemic has differed across continents. We hypothesized that regional differences in SARS-CoV-2 immunity might explain this observation. We therefore established the WWW Consortium in Ghana, W Africa; Jamaica, W Indies; and W London. Here, we describe the extent to which antibody immunity differs between these geographic locations.

*Methods:* The WWW Consortium harmonises across the HERITAGE (Accra, Ghana), WINDFall (Kingston, Jamaica) and Legacy (London, UK) studies, establishing sharing frameworks for samples, metadata, and data; related permissions and oversight; and associated physical and cloud infrastructure. With centralised testing, we performed serological assessments across all three locations at two snapshots in 2024 (April 1^st^ – August 18^th^; August 19^th^ – December 31^st^) using high-throughput live virus neutralization and anti-nucleocapsid IgG, including n=763 individuals.

**Findings:** We found that across all sites most participants had detectable neutralising antibody titres against JN.1 and XEC – the predominant variants in 2024. There were site-related differences in immunity: vaccine-included SARS-CoV-2 strains were better neutralised by participants from the Legacy study – Ancestral, BA.5, XBB.1.5 initially, and JN.1 after a homologous booster in autumn 2024. For HERITAGE, neutralisation of both alpha- (HCoV-229E) and beta-coronaviruses (HCoV-OC43) was higher than WINDFall suggesting a cross-coronavirus serological response in West Africa. Finally, antigenic cartography identified two distinct antibody landscapes, with JN.1 and XEC antigenically distant in Legacy, but not in HERITAGE and WINDFall.

**Interpretation:** There is international heterogeneity in SARS-CoV-2 antibody immunity. Global recommendations for vaccine strain selection should incorporate data from diverse populations to ensure accurate, equitable recommendations.

**Funding:** The Wellcome Trust.

## Introduction

Global endemicity of SARS-CoV-2 presents myriad challenges to individuals, to healthcare, and to wider society. Two challenges are directly informed by the study of serological immunity. The first is the decision to vaccinate further: when to recommend vaccination in at-risk populations, with which antigen(s), who is “at-risk”, and how to define and operationalize “at-risk”. The second challenge is the early – but accurate – declaration of a true immune escape variant, to enable prompt and proportionate proactive mitigations. To meet these challenges – particularly for worldwide vaccine antigen selection – requires representative serological snapshots from populations around the globe.

The global immune landscape against SARS-CoV-2 is shaped by the accumulation of sequential responses to vaccination and infection, themselves modulated at the individual-level by genes, environments and their interaction(s). Vaccine responses differ between people of different ethnicities. For example, mRNA vaccine responses in Australian First Nations were attenuated in the presence of chronic conditions, and comparable to non-indigenous participants in the absence of comorbidities (1). A meta-analysis of conjugate pneumococcal vaccination found geographic variation with stronger responses in the Western Pacific Region than Europe (2). Live attenuated vaccine antibody responses (measles IgG after MMR) differed between ethnicities within the USA (3). HLA polymorphisms will explain some of these differences (4), other variants in non-HLA, immunomodulatory genes such as IL-10 (5) will likely explain further differences, alongside non-genetic factors. Furthermore, responses to SARS-CoV-2 infection also show variability across ethnicities. In 2020, case fatality rates from COVID-19 in China were estimated to be 1·38% (95% credible interval 1·23–1·53) (6), albeit with a strong effect of age. Early in the UK’s experience, Black, Asian and Minority Ethnic (BAME) individuals were at increased risk of death after SARS-CoV-2 infection (7). This difference was not found in later waves (8), arguing against an unchanging genetic predisposition. Whereas several cities of Asia and Europe came close to healthcare systems failure (inadequate oxygen supply, insufficient ventilator capacity), this pattern was not observed across Africa despite – very generally – less healthcare provision (9). These differential responses to infection might be specific to SARS-CoV-2, be present across all *coronaviridae*, or reflect general sensitivity to antigen – perhaps at the risk of autoimmunity (10).

Here, we describe the cumulative impact of both vaccination and infection on SARS-CoV-2 antibody immunity at two timepoints in 2024 across three cities, on three continents: Accra, Ghana; Kingston, Jamaica and London, UK. We aligned three longitudinal cohort studies: in Accra, the HERITAGE study (11); in Kingston, the WINDFall study (12); and in London, the Legacy study (13–15). These snapshots simulate the data available for strain selection decisions, and for decisions to declare emergency or proactive measures against an emerging VOC which could be highly pathogenic. Next, we explore whether matching for age, sex and vaccine history removes the geographic heterogeneity. We consider whether pan-*coronaviridae* serological immunity might underlie these site-specific differences. Finally, we examine if antigenic cartography, a leading strain selection methodology, reflects the diversity of our consortium.

## Methods

### Ethics

The HERITAGE study protocol was reviewed and approved by the Ethics Review Committee of the Ghana Health Service (Reference: GHS-ERC-005/06/20). The Legacy study was approved by London Camden and Kings Cross Health Research Authority (HRA) Research and Ethics committee (REC, reference 20/HRA/4717) IRAS number 286469 and sponsored by University College London. The WINDFall study was approved by the Mona Campus Research Ethics Committee (reference CREC-MN.0250 2022/2023) with extensions granted yearly. All participants gave written informed consent for inclusion in the studies.

### Live virus microneutralisation assays

#### SARS-CoV-2

High-throughput live-virus microneutralisation assays for serum samples were performed as previously described (13–16). To ensure reproducibility/utility/comparability, assays were conducted against internal reference standards on each plate, calibrated to the WHO International Standard for anti-SARS-CoV-2 antibody (17,18).

#### HCoV-OC43 & HCoV-229E

High-throughput live virus microneutralisation assays were conducted adapting our existing assay platform for SARS-CoV-2 and influenza (13–16,19,20). Briefly, Mv1Lu and Huh-7 cells at 90–100% confluence were infected with HCoV-OC43 and HCoV-229E respectively in 384-well plates, together with serial dilutions of human sera. Infected plates were incubated at 33C for 24 hours. Post infection, cells were fixed with 4% formaldehyde, permeabilised using 0.2% Triton X-100 and 3% BSA in PBS, then stained with a virus-specific anti-N antibody (Sino Biological 40643-T62 for OC43 and Sino Biological 40640-T62 for 229E) followed by secondary staining using an Alexa Fluor 488 donkey anti-Rabbit antibody (Thermo A21206). Nuclear DNA of cells were stained using DAPI. Whole-well imaging at 5x magnification was performed using an Opera Phenix system (Revvity), and fluorescence metrics were quantified with Harmony software. Inhibition was determined by calculating the proportion of infected cell area (Anti-N positive) relative to the total cell area (DAPI positive). Serum neutralisation curves were generated by fitting a 4-parameter dose-response model in SciPy. Neutralising antibody titres (IC_50_) indicate the serum dilution required to reduce viral infection by 50% and are categorized as above the quantitative detection limit (complete neutralisation), within a qualitative range (partial neutralisation without a reliable dose-response fit), or showing weak or no detectable inhibition.

A pool of human sera was generated and characterised prior to usage as an internal reference standard on each assay plate.

### Roche anti-N

Anti-nucleocapsid IgG (anti-N IgG) detection Anti-nucleocapsid IgG was measured using the Elecsys Anti-SARS-COV-2 assay (Roche; 09203095190) run on a Cobas e411 analyser (Roche) in accordance with the manufacturer’s instructions. Serum was used for this immunoassay and results reported as reactive (positive) or non-reactive (negative).

### Data transfer, aggregation and analysis

Legacy study (NCT04750356) and WINDFall study data were collected and managed using REDCap electronic data capture tools hosted at University College London and The University of the West Indies, respectively (21,22). HERITAGE study data were collected as described (11). Pseudonymised participant data were transferred via secure file transfer protocols into a Trusted Research Environment (TRE) hosted on a Snowflake instance, which enforced role-based access controls, multi-factor authentication, and end-to-end encryption.

Data were imported into R from Snowflake prior to aggregation and analysis. Data were aggregated into a tidy-like data structure using R package *chronogram* (23). Data were manipulated, analysed and visualised using *tidyverse* R packages (24) including *dplyr* and *ggplot2*. Summary descriptions of the study cohorts were generated using *gtsummary*. Continuous data were reported as the median value and interquartile range (IQR) or the first and third quartiles (Q1; Q3).

Two group comparisons of neutralising antibody titres were performed with unpaired two-tailed Wilcoxon rank-sum tests. Statistical tests were conducted with the *rstatix* R package (25). Fold changes (FC) were estimated between groups with a 95% confidence interval (CI) with the *boot* R package using 1000 bootstrap resamples.

The *Racmacs* R package was used to perform antigenic cartography by importing neutralization-titre data into R and constructing antigen-serum maps. Quality control included assessing residuals by comparing fitted and measured titres and ensuring that the overall stress metric remained <0.2 (26). To evaluate map robustness, 100 bootstrap replicates were generated and visualized as uncertainty “blobs” around each antigen and serum point.

## Results

### The West Africa, West Indies, West London consortium

The West Africa, West Indies, West London consortium (WWWc) has recruiting cities across three separate continents (Africa, North America & Europe) **(Figure 1A)**. In Accra, Ghana, the HERITAGE study is a prospective cohort study within the Greater Accra Region of Ghana, Ghana’s most densely populated region (11). The WINDFall study is a prospective cohort study based in Kingston, Jamaica (12), and in London, UK, the Legacy study is a prospective cohort study conducted as a partnership between UCLH Biomedical Research Centre and the Francis Crick Institute enrolling both laboratory staff and healthcare workers (13–15). Using World Health Organisation data, confirmed cases in each country follow similar patterns, with a large Omicron BA.1 wave in all three nations, with smaller Delta and Alpha waves (**Figures 1B & C**). In Ghana and Jamaica, centralized testing was rapidly commenced, with a wave of confirmed cases pre-Alpha. Across all three nations, these confirmed cases will under-report true incidence due to the differential availability of testing, and this under-reporting was confirmed by serosurveys in Kingston (4.7% of persons reported previous COVID-19, yet 69% were positive for nucleocapsid IgG) (27). Whilst vaccination was offered in all three nations, the dates of vaccine deployment differed, with first doses in HERITAGE occurring approximately 1 year after WINDFall and Legacy, and HERITAGE following a short, 3 week-dose interval compared to longer intervals in WINDFall and Legacy (**Figure 1D**). Here, we focus on two aligned periods of synchronized study visits during 2024, comparing across these three studies between April 1st – August 18^th^ (*Period 1*, hereafter; **Table 1**), and between August 19^th^ – December 31^st^ (*Period 2*, hereafter; **Table 2**). For both periods, we report live virus microneutralisation titres against a suite of coronaviridae family members: SARS-CoV-2 and its variants, and common-cold causing betacoronavirus (HCoV-OC43) alphacoronavirus (HCoV-229E). Overall, 93% (508/545) and 95% (513/540) of participants had evidence of prior infection as their last anti-nucleocapsid IgG were positive in Period 1 or 2 respectively.

**Figure 1.**
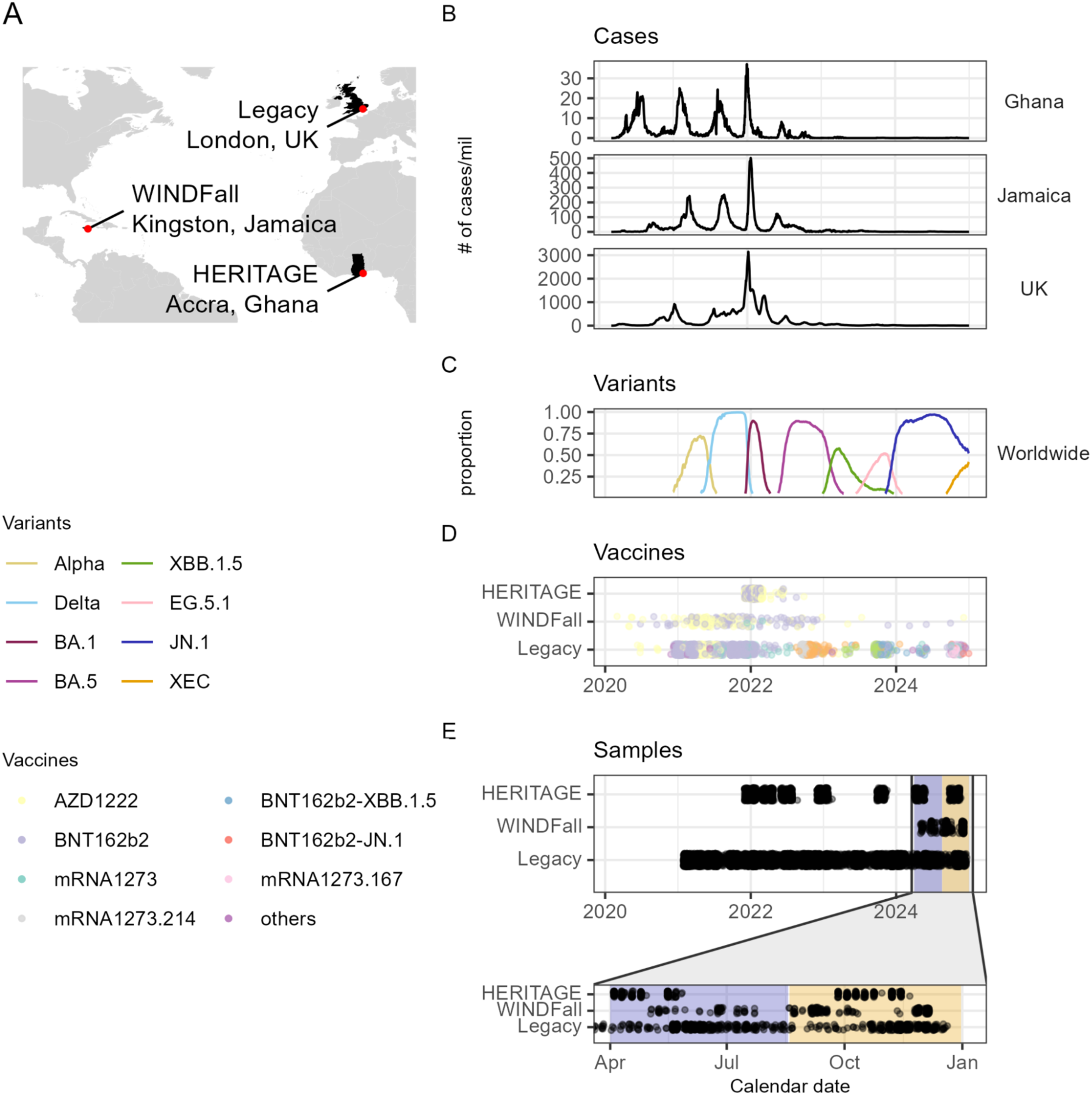
The West Africa, West Indies, West London consortium in 2024. **(A)** Map showing the 3 contributing studies to WWWc. **(B)** Numbers of daily new confirmed COVID-19 cases per million population over 2024-25 (World Health Organization (2025); Population based on various sources (2024) – with major processing by Our World in Data). Retrieval date: 5 June 2025 **(C)** The proportion of SARS-CoV-2 sequences corresponding to each of the main variants worldwide. Data from GISAID, with analysis from COVspectrum. Retrieval date: 6 June 2025 **(D)** Vaccines within WWWc are shown for HERITAGE, *WINDF*all, and Legacy **(E)** Serum samples used in this study, splitting 2024 into cohorts *Period 1* (April 1^st^ – August 18^th^) and *Period 2* (August 19^th^ – December 31^st^). See Tables 1 & 2 for demographic information for *Period 1* and *Period 2* respectively. Vaccine nomenclature: AZD1222 (ChAdOx-1, Oxford/AstraZeneca; adenoviral vector), BNT162b2 (Pfizer/BioNTech; mRNA vector), mRNA1273 (Moderna; mRNA vector), mRNA1273.214 (Moderna bivalent Ancestral & BA.1; mRNA vector), BNT162b2-XBB.1.5 (Pfizer/BioNTech monovalent XBB.1.5; mRNA vector), BNT162b2-JN.1 (Pfizer/BioNTech monovalent JN.1; mRNA vector), mRNA1273.167 (Moderna monovalent JN.1; mRNA vector).

**Table 1.**
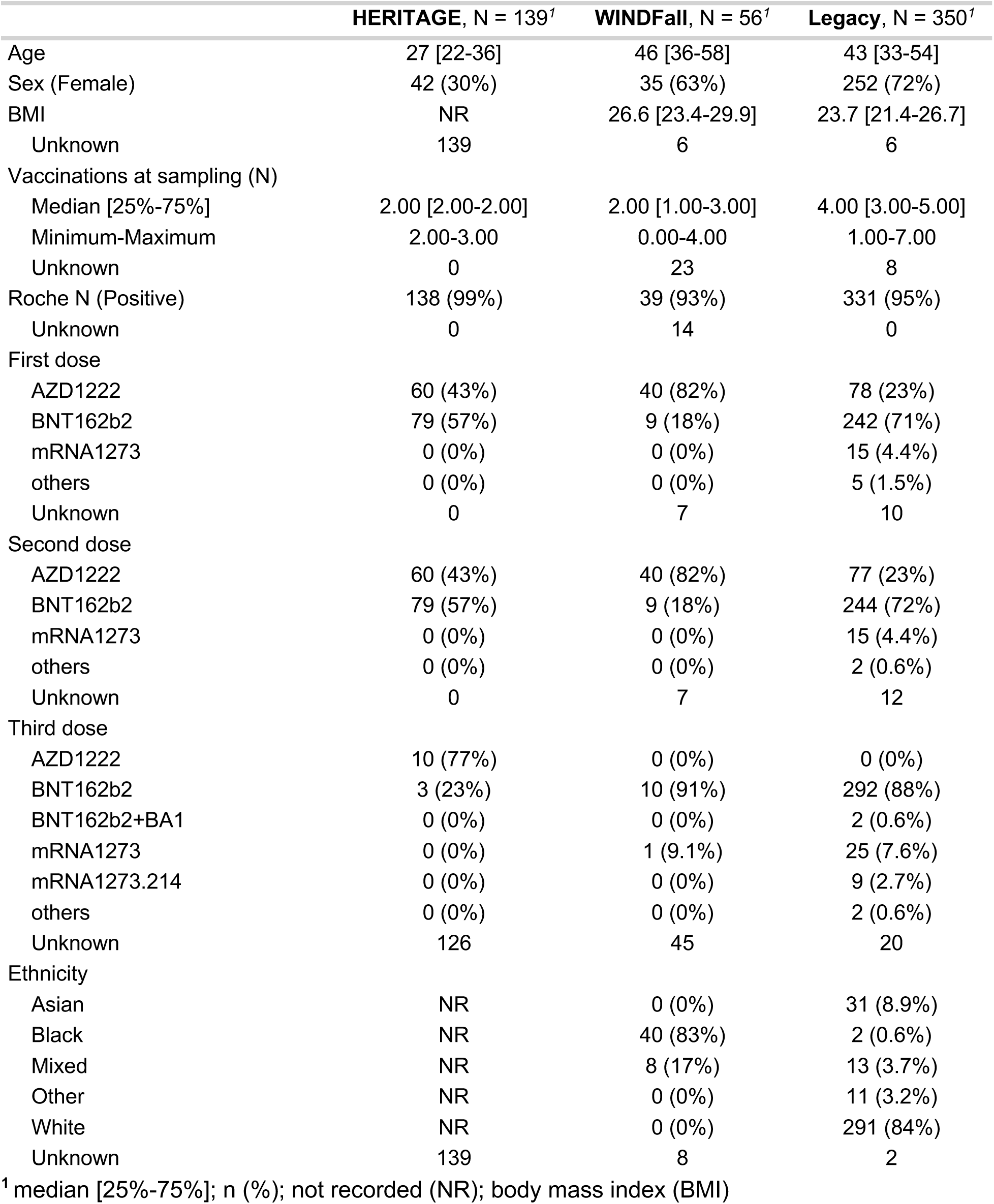
Participant demographics for Period 1 (P1, April 1^st^ – August 18^th^ 2024)

|  | HERITAGE, N = 139<sup>1</sup> | WINDFall, N = 56<sup>1</sup> | Legacy, N = 350<sup>1</sup> |
| --- | --- | --- | --- |
| Age | 27 [22-36] | 46 [36-58] | 43 [33-54] |
| Sex (Female) | 42 (30%) | 35 (63%) | 252 (72%) |
| BMI | NR | 26.6 [23.4-29.9] | 23.7 [21.4-26.7] |
| Unknown | 139 | 6 | 6 |
| Vaccinations at sampling (N) |  |  |  |
| Median [25%-75%] | 2.00 [2.00-2.00] | 2.00 [1.00-3.00] | 4.00 [3.00-5.00] |
| Minimum-Maximum | 2.00-3.00 | 0.00-4.00 | 1.00-7.00 |
| Unknown | 0 | 23 | 8 |
| Roche N (Positive) | 138 (99%) | 39 (93%) | 331 (95%) |
| Unknown | 0 | 14 | 0 |
| First dose |  |  |  |
| AZD1222 | 60 (43%) | 40 (82%) | 78 (23%) |
| BNT162b2 | 79 (57%) | 9 (18%) | 242 (71%) |
| mRNA1273 | 0 (0%) | 0 (0%) | 15 (4.4%) |
| others | 0 (0%) | 0 (0%) | 5 (1.5%) |
| Unknown | 0 | 7 | 10 |
| Second dose |  |  |  |
| AZD1222 | 60 (43%) | 40 (82%) | 77 (23%) |
| BNT162b2 | 79 (57%) | 9 (18%) | 244 (72%) |
| mRNA1273 | 0 (0%) | 0 (0%) | 15 (4.4%) |
| others | 0 (0%) | 0 (0%) | 2 (0.6%) |
| Unknown | 0 | 7 | 12 |
| Third dose |  |  |  |
| AZD1222 | 10 (77%) | 0 (0%) | 0 (0%) |
| BNT162b2 | 3 (23%) | 10 (91%) | 292 (88%) |
| BNT162b2+BA1 | 0 (0%) | 0 (0%) | 2 (0.6%) |
| mRNA1273 | 0 (0%) | 1 (9.1%) | 25 (7.6%) |
| mRNA1273.214 | 0 (0%) | 0 (0%) | 9 (2.7%) |
| others | 0 (0%) | 0 (0%) | 2 (0.6%) |
| Unknown | 126 | 45 | 20 |
| Ethnicity |  |  |  |
| Asian | NR | 0 (0%) | 31 (8.9%) |
| Black | NR | 40 (83%) | 2 (0.6%) |
| Mixed | NR | 8 (17%) | 13 (3.7%) |
| Other | NR | 0 (0%) | 11 (3.2%) |
| White | NR | 0 (0%) | 291 (84%) |
| Unknown | 139 | 8 | 2 |

**Table 2 –.** Participant demographics for Period 2 (P2, August 19^th^ – December 31^st^ 2024)

|  | HERITAGE, N = 139<sup>1</sup> | WINDFall, N = 146<sup>1</sup> | Legacy, N = 255<sup>1</sup> |
| --- | --- | --- | --- |
| Age | 27 [23-36] | 23 [20-35] | 47 [34-56] |
| Sex (Female) | 42 (30%) | 90 (62%) | 187 (73%) |
| BMI | NR | 26.0 [22.4-30.2] | 24.2 [21.7-27.4] |
| Unknown | 139 | 13 | 1 |
| Vaccinations at sampling (N) |  |  |  |
| Median [25%-75%] | 2.00 [2.00-2.00] | 1.00 [0.00-2.00] | 4.00 [3.00-6.00] |
| Minimum-Maximum | 2.00-3.00 | 0.00-4.00 | 1.00-9.00 |
| Unknown | 0 | 44 | 4 |
| Roche N (Positive) | 138 (99%)<sup>2</sup> | 131 (98%) | 244 (96%) |
| Unknown | 0 | 13 | 0 |
| First dose |  |  |  |
| AZD1222 | 63 (45%) | 39 (40%) | 48 (19%) |
| BNT162b2 | 76 (55%) | 58 (59%) | 189 (76%) |
| mRNA1273 | 0 (0%) | 1 (1.0%) | 7 (2.8%) |
| others | 0 (0%) | 0 (0%) | 4 (1.6%) |
| Unknown | 0 | 48 | 7 |
| Second dose |  |  |  |
| AZD1222 | 63 (45%) | 39 (40%) | 49 (20%) |
| BNT162b2 | 76 (55%) | 58 (59%) | 188 (76%) |
| mRNA1273 | 0 (0%) | 1 (1.0%) | 9 (3.6%) |
| others | 0 (0%) | 0 (0%) | 1 (0.4%) |
| Unknown | 0 | 48 | 8 |
| Third dose |  |  |  |
| AZD1222 | 10 (77%) | 0 (0%) | 0 (0%) |
| BNT162b2 | 3 (23%) | 5 (100%) | 211 (86%) |
| BNT162b2-JN.1 | 0 (0%) | 0 (0%) | 1 (0.4%) |
| BNT162b2+BA1 | 0 (0%) | 0 (0%) | 3 (1.2%) |
| mRNA1273 | 0 (0%) | 0 (0%) | 17 (7.0%) |
| mRNA1273.214 | 0 (0%) | 0 (0%) | 11 (4.5%) |
| others | 0 (0%) | 0 (0%) | 1 (0.4%) |
| Unknown | 126 | 141 | 11 |
| Ethnicity |  |  |  |
| Asian | NR | 1 (0.7%) | 25 (9.8%) |
| Black | NR | 112 (80%) | 4 (1.6%) |
| Mixed | NR | 27 (19%) | 10 (3.9%) |
| Other | NR | 0 (0%) | 6 (2.4%) |
| White | NR | 0 (0%) | 209 (82%) |
| Unknown | 139 | 6 | 1 |

<sup>1</sup> median [25%-75%]; n (%); not recorded (NR); body mass index (BMI)
<sup>2</sup> last available anti-N IgG result. For HERITAGE this is Period 1.

### SARS-CoV-2 humoral immunity in the West Africa, West Indies, West London consortium in 2024

To test for serological heterogeneity between these three continents, we measured serum neutralisation titres against SARS-CoV-2 at synchronized study visits across all three sites through 2024 between April 1st – August 18^th^ (*Period 1*) and August 19^th^ – December 31^st^ (*Period 2*) **(Figures 2A & B)**. As part of synchronization, we ensured that an individual was present only once in Period 1 or Period 2, retaining the most recent visit. In both Period 1 and Period 2, quantifiable titres (IC_50_>40) against Ancestral, BA.2, and JN.1 were observed (*Period 1*: HERITAGE 139/139; WINDFall 56/56; Legacy 350/350 & *Period 2*: HERITAGE 139/139; WINDFall146/146; Legacy 255/255). XEC, first detected August 7^th^ 2024 (28), was neutralized – with quantifiable titres – by over 90% of participants in Period 1 (HERITAGE 139/139; WINDFall 53/56; Legacy 325/350) subsequently rising to over 97% in Period 2 as global prevalence increased (HERITAGE 139/139; WINDFall144/146; Legacy 248/255) (29). Median titres against Ancestral, BA.2, and BA.5 and XBB.1.5 were consistently higher in Legacy participants than their counterparts in HERITAGE and WINDFall. Neutralization of Ancestral was 3-fold higher compared to HERITAGE in Period 1 (FC=3.0; CI 2.1-4.3) and Period 2 (FC=3.0; CI 2.3-4.3) (**Table 3**). When compared to WINDFall, Ancestral neutralization in Legacy was between 1.5-4-fold higher in Period 1 (FC=1.5; CI 1.2-2.1) and Period 2 (FC=3.8; CI 2.5-5.1).

**Figure 2.**
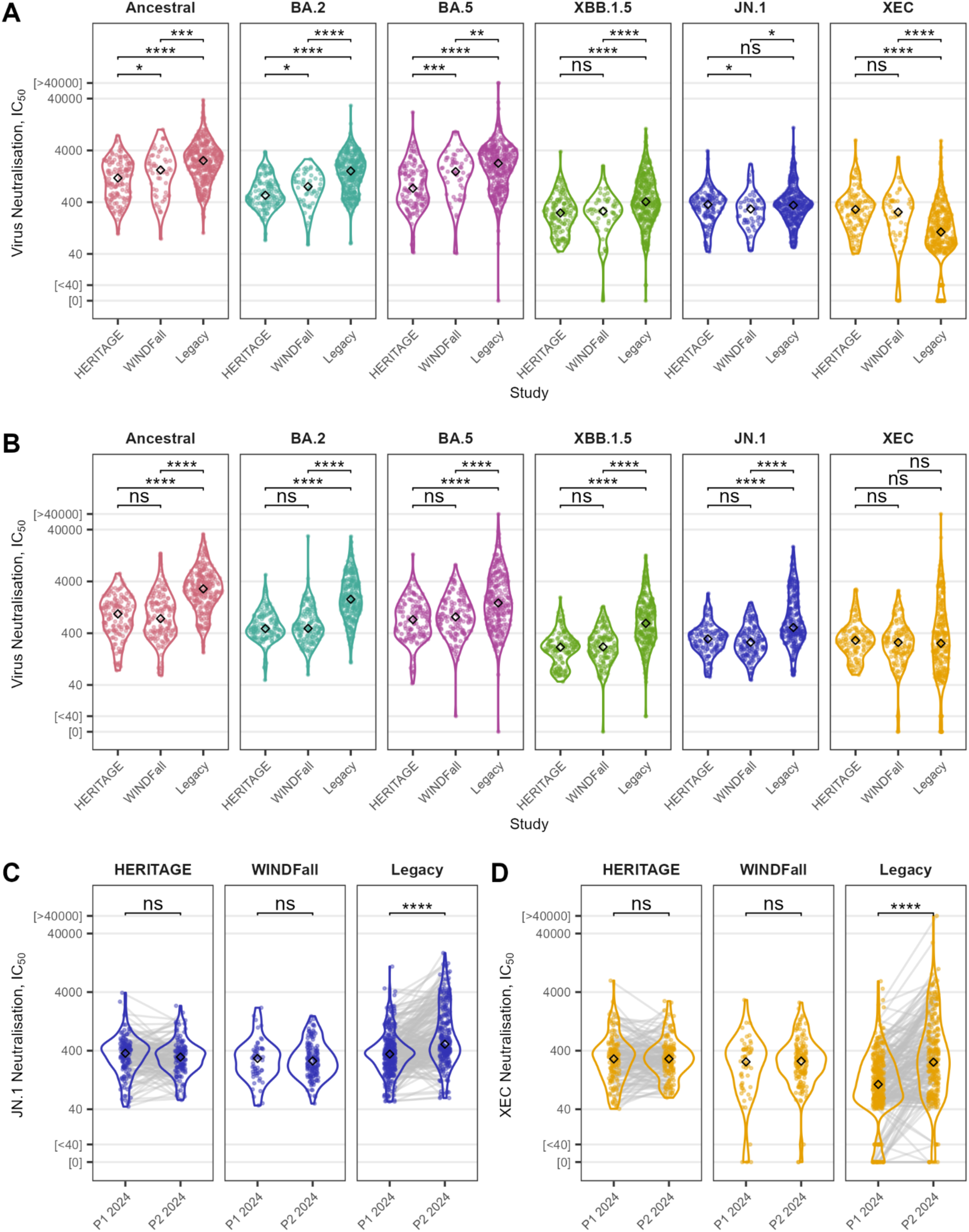
Continental heterogeneity in SARS-CoV-2 serological immunity during 2024. Serum neutralisation titres against SARS-CoV-2 variants from HERITAGE, WINDFall, and Legacy participants at two time points in 2024: (**A**) Period 1, *P1*, April 1^st^ – August 18^th^; and (**B**) Period 2, *P2*, August 19^th^ – December 31^st^. Titres are compared between time points for JN.1 (**C**) and XEC (**D**). Titres are plotted as Log_2_-transformed 50% inhibitory concentrations (IC_50_), the reciprocal of the serum dilution at which 50% of viral infection is inhibited. Significance was determined by two-tailed Wilcoxon rank-sum tests and is indicated by asterisks: * p < 0.05; ** p < 0.01; *** p < 0.001; **** P < 0.0001; ns, not significant (p ≥ 0.05).

**Table 3.** Fold change (FC) in serum neutralisation titres between studies by timepoint (P1, April 1^st^ – August 18^th^ 2024; P2, August 19^th^ – December 31^st^ 2024)

| Time point | Variant | Group 1 | Group 2 | FC | Lower | Upper | P |
| --- | --- | --- | --- | --- | --- | --- | --- |
| P1 | Ancestral | HERITAGE | WINDFall | 1.4 | 0.9 | 2.2 | 0.041 |
| P1 | BA.2 | HERITAGE | WINDFall | 1.5 | 1.1 | 2 | 0.04 |
| P1 | BA.5 | HERITAGE | WINDFall | 2.1 | 1.3 | 3.1 | 0.001 |
| P1 | XBB.1.5 | HERITAGE | WINDFall | 1.1 | 0.9 | 1.3 | 0.658 |
| P1 | JN.1 | HERITAGE | WINDFall | 0.8 | 0.6 | 1.1 | 0.048 |
| P1 | XEC | HERITAGE | WINDFall | 0.9 | 0.6 | 1.3 | 0.29 |
| P1 | Ancestral | HERITAGE | Legacy | 2.2 | 1.6 | 3 | 0 |
| P1 | BA.2 | HERITAGE | Legacy | 2.9 | 2.4 | 3.5 | 0 |
| P1 | BA.5 | HERITAGE | Legacy | 3 | 2.1 | 4.3 | 0 |
| P1 | XBB.1.5 | HERITAGE | Legacy | 1.6 | 1.4 | 2 | 0 |
| P1 | JN.1 | HERITAGE | Legacy | 1 | 0.8 | 1.1 | 0.875 |
| P1 | XEC | HERITAGE | Legacy | 0.4 | 0.3 | 0.5 | 0 |
| P1 | Ancestral | WINDFall | Legacy | 1.5 | 1.2 | 2.1 | 0.001 |
| P1 | BA.2 | WINDFall | Legacy | 2 | 1.5 | 2.6 | 0 |
| P1 | BA.5 | WINDFall | Legacy | 1.5 | 1.1 | 2.1 | 0.008 |
| P1 | XBB.1.5 | WINDFall | Legacy | 1.5 | 1.2 | 1.8 | 0 |
| P1 | JN.1 | WINDFall | Legacy | 1.2 | 0.9 | 1.7 | 0.023 |
| P1 | XEC | WINDFall | Legacy | 0.4 | 0.3 | 0.7 | 0 |
| P2 | Ancestral | HERITAGE | WINDFall | 0.8 | 0.5 | 1.3 | 0.456 |
| P2 | BA.2 | HERITAGE | WINDFall | 1 | 0.9 | 1.2 | 0.762 |
| P2 | BA.5 | HERITAGE | WINDFall | 1.1 | 0.9 | 1.6 | 0.126 |
| P2 | XBB.1.5 | HERITAGE | WINDFall | 1 | 0.8 | 1.3 | 0.319 |
| P2 | JN.1 | HERITAGE | WINDFall | 0.9 | 0.7 | 1.1 | 0.144 |
| P2 | XEC | HERITAGE | WINDFall | 0.9 | 0.7 | 1.1 | 0.261 |
| P2 | Ancestral | HERITAGE | Legacy | 3 | 2.3 | 4.3 | 0 |
| P2 | BA.2 | HERITAGE | Legacy | 3.7 | 3.1 | 4.4 | 0 |
| P2 | BA.5 | HERITAGE | Legacy | 2.1 | 1.6 | 2.9 | 0 |
| P2 | XBB.1.5 | HERITAGE | Legacy | 2.9 | 2.2 | 3.7 | 0 |
| P2 | JN.1 | HERITAGE | Legacy | 1.7 | 1.3 | 2.1 | 0 |
| P2 | XEC | HERITAGE | Legacy | 0.9 | 0.7 | 1.1 | 0.4 |
| P2 | Ancestral | WINDFall | Legacy | 3.8 | 2.5 | 5.1 | 0 |
| P2 | BA.2 | WINDFall | Legacy | 3.7 | 3.1 | 4.4 | 0 |
| P2 | BA.5 | WINDFall | Legacy | 1.9 | 1.4 | 2.3 | 0 |
| P2 | XBB.1.5 | WINDFall | Legacy | 2.9 | 2.1 | 3.8 | 0 |
| P2 | JN.1 | WINDFall | Legacy | 1.9 | 1.5 | 2.4 | 0 |
| P2 | XEC | WINDFall | Legacy | 1 | 0.7 | 1.3 | 0.832 |

The WHO Technical Advisory Group on COVID-19 Vaccine Composition (TAG-CO-VAC) has recommended that manufacturers continue the use of monovalent JN.1 lineage vaccine antigen in vaccine development in 2025, matching the antigen used in 2024 (30). During Period 1 2024, median titres against JN.1 were comparable in both HERITAGE (IC_50_=361.2) and Legacy (IC_50_=349.4) participants and were marginally lower in WINDFall (IC_50_=294.7; P=0.02). However, when participants were sampled later in 2024 the titres were almost 2-fold higher in Legacy compared to Heritage (FC=1.7; CI 1.3-2.1) and WINDFall (FC=1.9; CI 1.5-2.4). Expectedly, comparing Period 1 to Period 2, Legacy showed a significant increase in JN.1 titres, reflecting the UK’s autumn 2024 JN.1 mRNA booster campaign **(Figure 2C**; **Supplemental Figure 1)**, and a cross-neutralisation boost against XEC **(Figure 2D)**. Neutralisation of the XEC variant in HERITAGE (FC=2.5; CI 2.0-3.3) and WINDFall (FC=2.5; CI 1.4-3.3) participants was double that observed in Legacy in Period 1 2024. By contrast, when sampled later in 2024 no significant differences in XEC immunity were apparent (P > 0.05), reflecting either JN.1 booster cross-protection against XEC, or additional infections in Legacy cohort.

### Age, sex and vaccine matched SARS-CoV-2 humoral immunity in the West Africa, West Indies, West London consortium in 2024

In addition to the geographic differences between participants of the three studies, the cohorts each differ substantially by age, sex and vaccination history. To test the impact these potential confounders have on our analysis, we identified sub-cohorts of individuals that were matched based on age group (≤35, 35-50, ≥50yo), sex, and the number of SARS-CoV-2 vaccinations (≥2 doses) during Period 2 (**Table 4**). The resulting selection was also matched in time between all three sites (**Figure 3A**).

**Figure 3.**
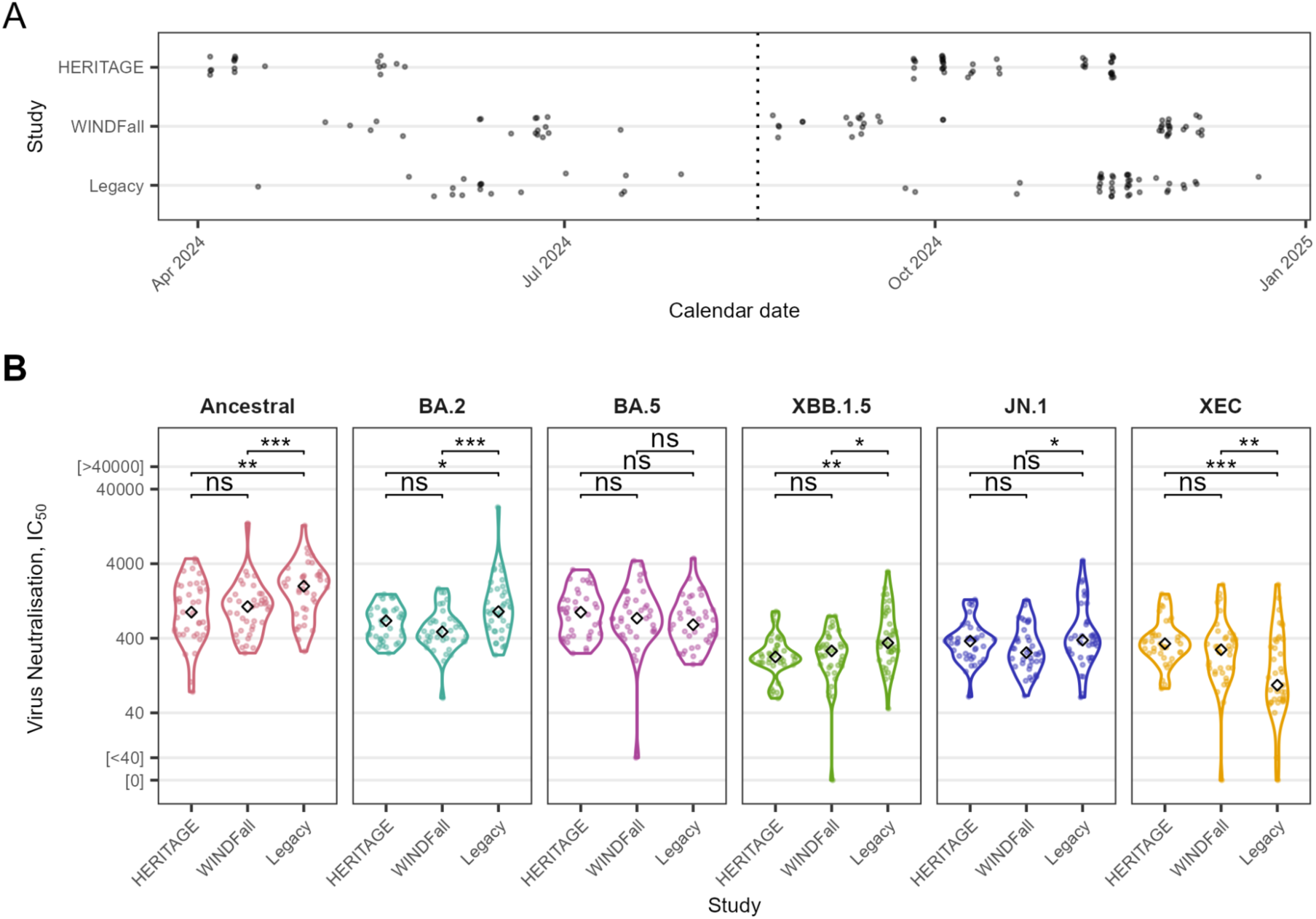
Spatial differences in antibody immunity in a matched West Africa, West Indies, West London sub-cohort. (**A**) Serum samples retained after matching, splitting 2024 into time points Period 1 (April 1st – August 18th) and Period 2 (August 19th – December 31st) indicated with the dashed line. See Table 4 & Supplemental Table 1 for demographic information for the matched Period 2 and Period 1, respectively. (**B**) Serum neutralisation titres in Period 2 2024 (August 19^th^ – December 31^st^) from HERITAGE, WINDFall, and Legacy participants matched based on age group (≤35, 35-50, ≥50), sex, and number of vaccinations (≥2 doses).Titres are plotted as Log_2_-transformed 50% inhibitory concentrations (IC_50_), the reciprocal of the serum dilution at which 50% of viral infection is inhibited. Significance was determined by two-tailed Wilcoxon rank-sum tests and is indicated by asterisks: * p < 0.05; ** p < 0.01; *** p < 0.001; **** P < 0.0001; ns, not significant (p ≥ 0.05).

**Table 4.** Participant demographics for Period 2 (P2, August 19th – December 31st 2024) when matched on age group (≤35, 35-50, ≥50), sex, and the number of SARS-CoV-2 vaccinations (≥2 doses & < 4 doses)

|  | HERITAGE, N = 39<sup>1</sup> | WINDFall, N = 39<sup>1</sup> | Legacy, N = 39<sup>1</sup> |
| --- | --- | --- | --- |
| Age | 30 [23-38] | 28 [21-38] | 31 [28-40] |
| Sex (Female) | 26 (67%) | 26 (67%) | 26 (67%) |
| BMI | NR | 27.2 [23.5-30.2] | 24.8 [22.1-26.5] |
| Unknown | 39 | 4 | 0 |
| Vaccinations at sampling (N) |  |  |  |
| Median [25%-75%] | 2.00 [2.00-2.50] | 2.00 [2.00-2.00] | 3.00 [3.00-3.00] |
| Minimum-Maximum | 2.00-3.00 | 2.00-3.00 | 2.00-3.00 |
| Roche N (Positive) | 0 (NA%) | 36 (100%) | 39 (100%) |
| Unknown | 39 | 3 | 0 |
| First dose |  |  |  |
| AZD1222 | 22 (56%) | 16 (42%) | 10 (26%) |
| BNT162b2 | 17 (44%) | 21 (55%) | 25 (64%) |
| mRNA1273 | 0 (0%) | 1 (2.6%) | 3 (7.7%) |
| others | 0 (0%) | 0 (0%) | 1 (2.6%) |
| Unknown | 0 | 1 | 0 |
| Second dose |  |  |  |
| AZD1222 | 22 (56%) | 16 (42%) | 11 (28%) |
| BNT162b2 | 17 (44%) | 21 (55%) | 25 (64%) |
| mRNA1273 | 0 (0%) | 1 (2.6%) | 3 (7.7%) |
| Unknown | 0 | 1 | 0 |
| Third dose |  |  |  |
| AZD1222 | 8 (80%) | 0 (NA%) | 0 (0%) |
| BNT162b2 | 2 (20%) | 0 (NA%) | 33 (87%) |
| BNT162b2-JN.1 | 0 (0%) | 0 (NA%) | 1 (2.6%) |
| mRNA1273 | 0 (0%) | 0 (NA%) | 4 (11%) |
| Unknown | 29 | 39 | 1 |
| Ethnicity |  |  |  |
| Asian | NR | 1 (2.6%) | 4 (10%) |
| Black | NR | 30 (79%) | 1 (2.6%) |
| Mixed | NR | 7 (18%) | 2 (5.1%) |
| Other | NR | 0 (0%) | 1 (2.6%) |
| White | NR | 0 (0%) | 31 (79%) |
| Unknown | 39 | 1 | 0 |

<sup>1</sup> median [25%-75%]; n (%); not recorded (NR); body mass index (BMI)

Titres against Ancestral and BA.2 remained lower in HERITAGE and WINDFall compared with Legacy (P<0.001) (**Figure 3B**). Conversely, Legacy participants exhibited only 20-30% of the XEC neutralisation observed in matched sera from HERITAGE (FC=0.3; CI 0.1-0.8) (**Table 5**) and WINDFall (FC=0.2; CI 0.1-0.5) in Period 2. Additionally, the increase in titres against JN.1 after a homologous booster was expectedly absent in this un-boosted Legacy sub-cohort, and consequently titres against JN.1 were comparable to HERITAGE and WINDFall. The matching procedure was repeated during Period 1 (**Supplemental Table 1**). Repeating the comparison re-iterated that XEC titres were lower in Legacy during Period 1 in a matched cohort (**Supplemental Figure 2**; **Supplemental Table 2**).

**Table 5.** Fold change (FC) in serum neutralisation titres in matched sub-cohorts during Period 2 (P2, August 19th – December 31st 2024)

| Time point | Variant | Group 1 | Group 2 | FC | Lower | Upper | P |
| --- | --- | --- | --- | --- | --- | --- | --- |
| P2 | Ancestral | HERITAGE | WINDFall | 1.2 | 0.6 | 2.2 | 0.9320 |
| P2 | BA.2 | HERITAGE | WINDFall | 0.7 | 0.5 | 1.1 | 0.0880 |
| P2 | BA.5 | HERITAGE | WINDFall | 0.8 | 0.6 | 1.3 | 0.9960 |
| P2 | XBB.1.5 | HERITAGE | WINDFall | 1.2 | 0.9 | 1.5 | 0.5830 |
| P2 | JN.1 | HERITAGE | WINDFall | 0.7 | 0.5 | 1.1 | 0.0690 |
| P2 | XEC | HERITAGE | WINDFall | 0.8 | 0.4 | 1.1 | 0.1170 |
| P2 | Ancestral | HERITAGE | Legacy | 2.2 | 1.3 | 4.1 | 0.0030 |
| P2 | BA.2 | HERITAGE | Legacy | 1.3 | 0.8 | 2.0 | 0.0220 |
| P2 | BA.5 | HERITAGE | Legacy | 0.7 | 0.5 | 1.1 | 0.1290 |
| P2 | XBB.1.5 | HERITAGE | Legacy | 1.5 | 1.2 | 2.2 | 0.0020 |
| P2 | JN.1 | HERITAGE | Legacy | 1.0 | 0.8 | 1.5 | 0.4690 |
| P2 | XEC | HERITAGE | Legacy | 0.3 | 0.2 | 0.7 | 0.0002 |
| P2 | Ancestral | WINDFall | Legacy | 1.9 | 1.3 | 2.8 | 0.0010 |
| P2 | BA.2 | WINDFall | Legacy | 1.9 | 1.1 | 2.5 | 0.0005 |
| P2 | BA.5 | WINDFall | Legacy | 0.8 | 0.5 | 1.3 | 0.1620 |
| P2 | XBB.1.5 | WINDFall | Legacy | 1.3 | 0.9 | 1.9 | 0.0150 |
| P2 | JN.1 | WINDFall | Legacy | 1.5 | 1.0 | 2.0 | 0.0270 |
| P2 | XEC | WINDFall | Legacy | 0.3 | 0.2 | 0.8 | 0.0070 |

### Pan-coronavirus neutralising immunity differs around the world

Having established that geographic location affected SARS-CoV-2 neutralisation, we next considered what aspect of geography might explain these differences. Here, we tested the hypothesis that pan-coronavirus immunity might differ across continents. Responses to seasonal human coronaviruses (HCoV) including OC43 and 229E were tested across the two sampling periods. In Period 1, 229E was more robustly neutralized by sera from HERITAGE compared to Legacy (FC 1.2; CI 1.1-1.3) and WINDFall (FC 1.3; CI 1-1.5) (**Figure 4A**, **Supplemental Table 3**). This effect was also apparent when tested in samples from Period 2 2024 (**Figure 4B**, **Supplemental Table 3**). The neutralization of OC43 was significantly higher in Period 1 HERITAGE samples compared to participants from Legacy (FC 1.1; CI 1.0-1.2) and WINDFall (FC 1.3; CI 1.1-1.5). Titres against OC43 were comparable with Legacy during Period 2 (P>0.05), but significantly higher than those of WINDFall (FC 1.6; CI 1.5-1.8).

**Figure 4.**
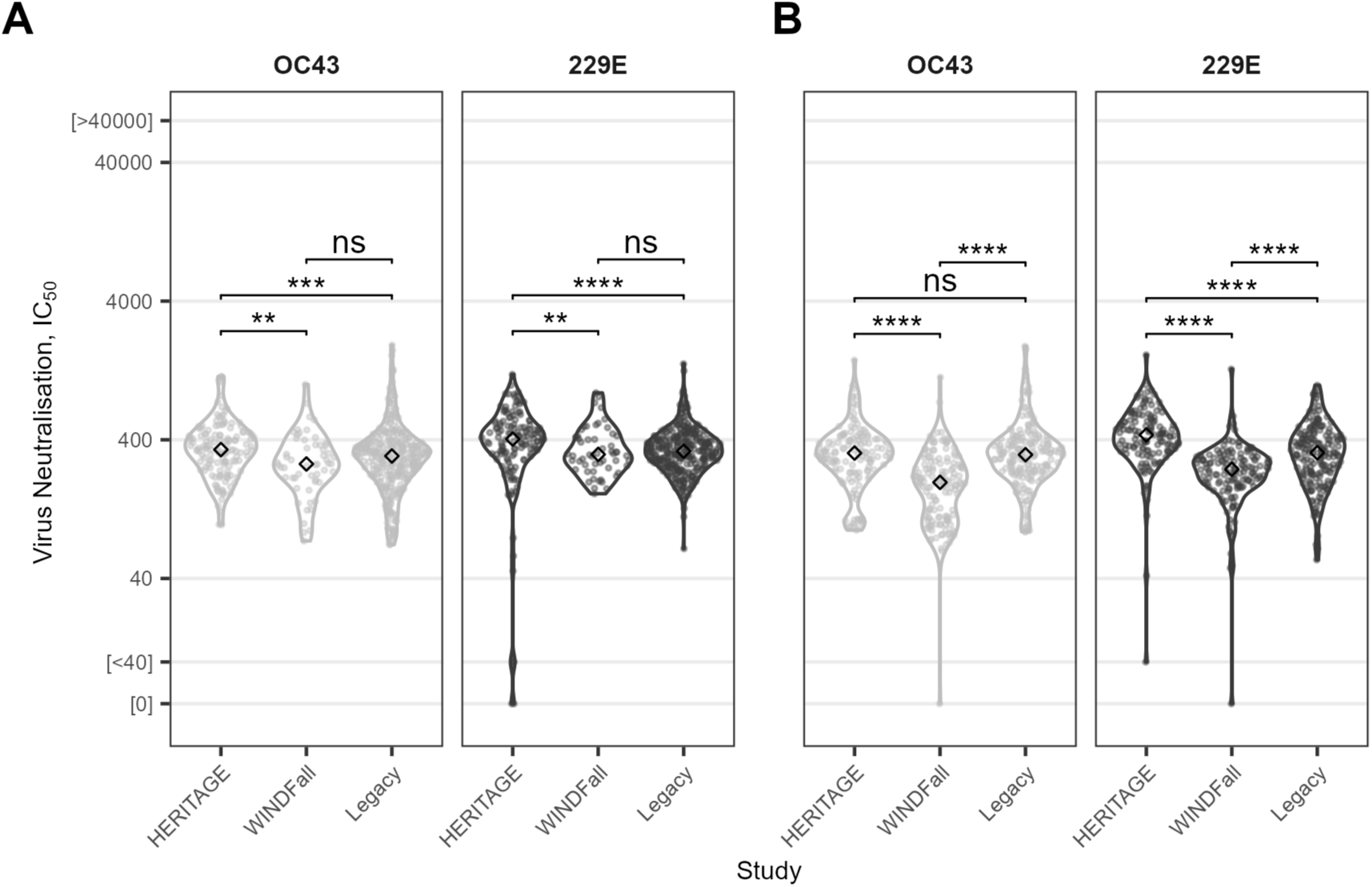
Common coronavirus neutralising immunity differs around the world. Serum neutralisation titres against human coronaviruses OC43 and 229E from HERITAGE, WINDFall, and Legacy participants at two timepoints in 2024: (**A**) Period 1, April 1^st^ – August 18^th^; and (**B**) Period 2, August 19^th^ – December 31^st^. Titres are plotted as Log_2_-transformed 50% inhibitory concentrations (IC_50_), the reciprocal of the serum dilution at which 50% of viral infection is inhibited. Significance was determined by two-tailed Wilcoxon rank-sum tests and is indicated by asterisks: * p < 0.05; ** p < 0.01; *** p < 0.001; **** P < 0.0001; ns, not significant (p ≥ 0.05).

### Antigenic cartography of SARS-CoV-2 antibody responses shows continental drift

To visualize the antigenic relationships between the SARS-CoV-2 variants and the sera and how this varied between study and time point, antigenic maps were generated (**Figure 5**). The resulting maps position each antigen and serum sample in antigenic space such that the spacing between grid lines correspond to 1 antigenic unit, or a two-fold dilution of anti-serum. These antigenic maps could be used to approximate the existing immune landscape, and to “place” new variants within that scene, and are currently used as a tool to guide vaccine recommendations (31).

**Figure 5.**
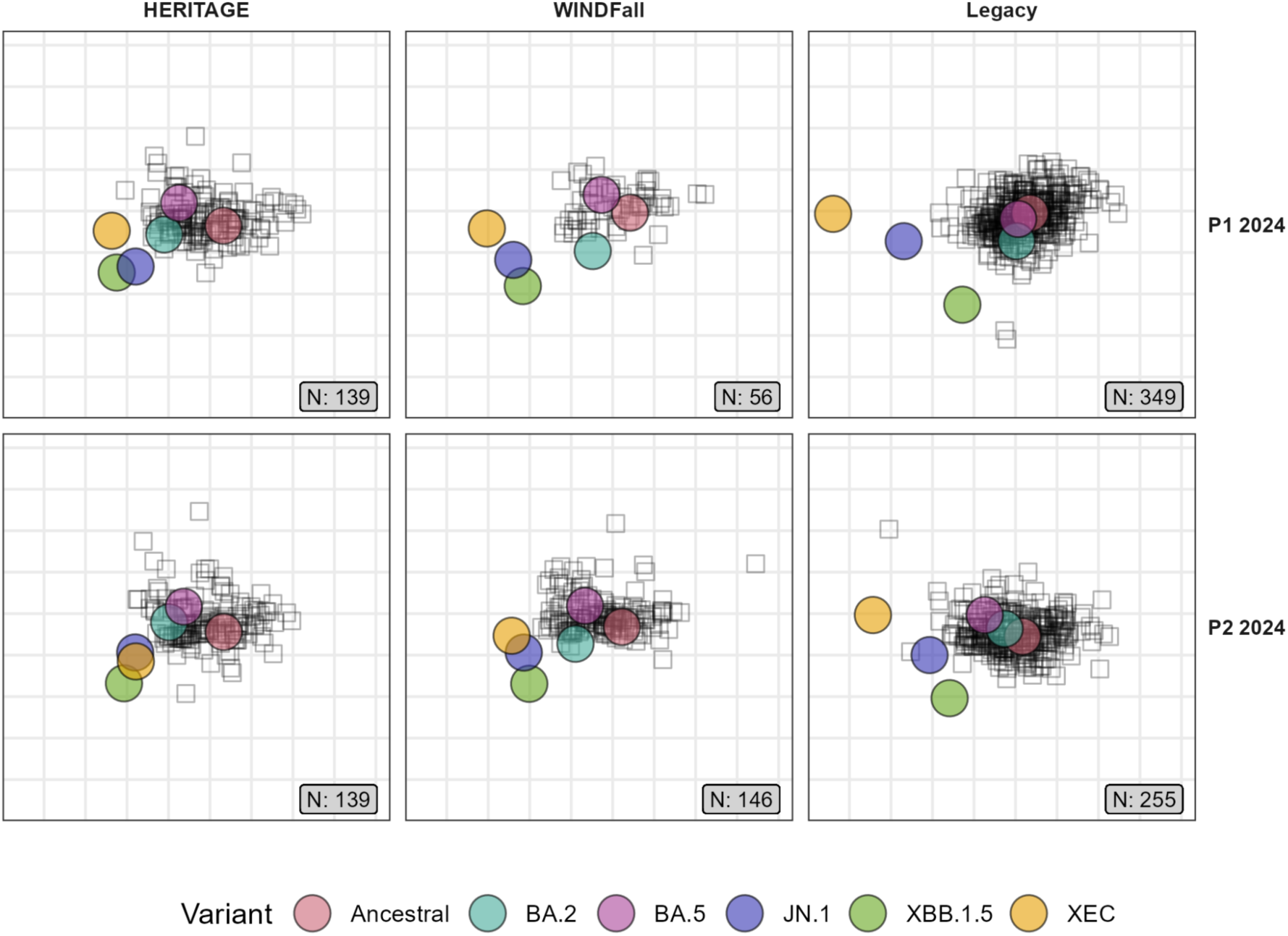
Geographic differences in antigenic space. Antigenic maps of SARS-CoV-2 variants for each study and timepoint in 2024, with columns HERITAGE, WINDFall, and Legacy (columns left to right) and rows Period 1 (top) and Period 2 (bottom). Coloured circles denote virus antigens; black squares indicate individual sera; the N labels show the count of sera. Maps were generated in Racmacs; one grid unit represents a two-fold change in neutralization titre. Proximity between a serum and a variant reflects stronger neutralisation of that variant.

A consistent pattern was observed across studies and time-point, whereby Ancestral and earlier Omicron variants BA.2 and BA.5 grouped together so are less antigenically distant. Conversely, XBB.1.5, JN.1 and XEC appear more distant relative to Ancestral. Moreover, the maps depict most sera close to the Ancestral, BA.2, BA.5 grouping of antigens, in line with the higher neutralisation titres observed against these variants **(Figures 2 & 3)**.

Where the three antigenic maps diverge is the relative placement of XEC, JN.1 and XBB.1.5 **(Figure 5)**. In Period 1, XEC is approximately 5 antigenic units from Ancestral in the map generated from UK-based Legacy samples. In contrast, XEC is only around 3 antigenic units from Ancestral in Jamaica-based WINDFall and 2 antigenic units in Accra-based HERITAGE. Furthermore, antigenic distance between XEC and JN.1 is highest in Legacy compared to the other two studies. This pattern is repeated when the maps were generated from samples in Period 2, with an approximate doubling of the XEC-ancestral antigenic distance in Legacy compared to HERITAGE and WINDFall (**Figure 5**). These cartographic differences at Period 1 and Period 2 are present when considering the age-, sex- and vaccine-history matching sub-cohorts (**Supplemental Figure 3).**

## Discussion

We described the design and initiation of the West Africa, West Indies, West London consortium, and report serological immunity against SARS-CoV-2 across 2024. We found differential patterns of immunity: the London cohort, in early 2024, had increased antibody titres against older variants that were either present in vaccines (Ancestral, BA.5) or highly related to those in a vaccine (BA.2 neutralisation, having received BA.1 containing bivalent). Conversely, participants in both Accra and Kingston demonstrated higher neutralising antibody titres against XEC in early 2024, a variant that did not dominate global sequencing until late 2024 **(Figures 2 & 3)**. Together, a possible explanation is that HERITAGE and WINDFall participants mount serological responses that in some regard forecasts future variants more successfully than in Legacy – perhaps through the generation of more antibody clones that are cross-reactive to new lineages, or the production of more antibody per cell from similar numbers of Spike-specific B cells that cross-neutralise. By examining neutralisation to two laboratory strains of common coronaviruses, as prototypical alphacoronavirus (229E) or betacoronavirus (OC43), we found that HERITAGE had broader pan-coronaviridae neutralisation than WINDFall or Legacy. Overall, this suggests that exposure to additional coronaviruses (across the family) might explain the breadth of responses in HERITAGE; and that another mechanism drives the XEC neutralisation in WINDFall, which tended to have lower OC43 and 229E neutralisation whilst maintaining XEC neutralisation.

We found that antigenic cartography is not homogeneous across WWWc **(Figure 5)**. Using antigenic cartography for vaccine strain selection, with global recommendations for manufacturers, is better performed with global study populations processed through a single assay, such that experimental variation is minimised allowing geography-related variation to be fully appreciated, and not overlook from obscuring per-assay effects, or compressed by cross-study normalisation.

Our study has some limitations. Firstly, as discussed above, continental-level trends in immunity risks an over-simplification of more nuanced differences and patterns. Secondly, whilst each study recruited locally, using local knowledge, we cannot guarantee that our study populations mirror the wider population from which they were drawn – for example Legacy is, due to its healthcare worker participants, more highly vaccinated than the London adult population. Thirdly, this work is retrospective, observational and non-randomised, and affected by differences in national policies and time that could all contribute to confounding. For example, perfect matching for total vaccinations is not possible: there was high uptake of a third dose offered in the UK at the start of the BA.1 wave, but lower uptake of additional doses in Ghana and Jamaica. However, for prospective decisions regarding strain selection or widespread re-vaccination, the current snapshot is critical, not the vaccination route taken to arrive at that snapshot.

In summary, we have demonstrated that SARS-CoV-2 antibody immunity differs across these three continents. Within this “location” are myriad potential determinants of antibody responses and unpicking these is the focus of the consortium’s ongoing and future work: we will consider HLA (4), non-HLA genetics (for example blood group (32)), malaria (33), previous virome exposures more broadly, and examine behavioural differences which might modulate immune status.

## Contributors

DG – data curation, formal analysis, software, visualisation, writing – original draft, writing review & editing

OH – data curation, investigation

EMKM – data curation, investigation

SAR – data curation, investigation

FS – project administration

JJA – conceptualisation, supervision, writing – review & editing.

GA – conceptualisation, supervision, writing – review & editing.

DLVB – conceptualisation, funding acquisition, supervision, writing – review & editing.

YB – conceptualisation, supervision, writing – review & editing.

EJC – conceptualisation, software, supervision, visualisation, writing – original draft, writing – review & editing.

CVFC – conceptualisation, supervision, writing – review & editing.

AK – conceptualisation, supervision, writing – review & editing.

PQ – conceptualisation, supervision, writing – review & editing.

ECW – conceptualisation, supervision, writing – review & editing.

MYW – conceptualisation, investigation, methodology, supervision, writing – review & editing.

All authors had final responsibility for the decision to submit for publication. DLVB had access to and verified the data reported in this manuscript.

## Data sharing statement

Requests to access de-identified participant data will first be considered by the WWWc Data Management Board (DMB), to which partners nominated a representative from their respective organisations (but outside their core team). After approval by the WWWc DMB, requests will be passed to the relevant study, or studies, to complete per study access request processes. Depending on the nature of the request Material Transfer Agreement(s) might be required, with one or more consortium studies, prior to data sharing.

## Acknowledgements

We thank all study participants in the HERITAGE, WINDFall and Legacy studies. We thank the study site clinical personnel who helped with recruitment of participants and study visits. We are grateful to the legal representatives of our institutions who worked hand-in-hand between different legal jurisdictions. We acknowledge the help of the Crick’s Information Technology Office, Emily Till, Davina Legah, Karen Ambrose who built and maintained the WWWc snowflake instance. We would like to thank the staff of the NIHR Clinical Research Facility at UCLH including Miguel Alvarez and Marivic Ricamara. We would like to thank the staff of the Scientific Technology Platforms (STPs) and COVID-19 testing pipeline at the Francis Crick Institute. We thank Prof. Alex Sigal, Dr Khadija Khan and Prof. Tulio de Oliveira of Africa Health Research Institute, Durban, South Africa, Dr Laura McCoy of UCL, and Prof. Gavin Screaton of the University of Oxford, for providing source material.

Shanice Redman and Jenene Cameron are global infectious diseases scholars who received mentored research training in work leading to the development of this manuscript. This training was supported in part by the University of Buffalo Clinical and Translational Science Institute (award no. UL1TR001412) and the Global Infectious Diseases Research Training Program (award no. D43TW010919).

This work was supported by the Wellcome Trust (226142/Z/22/Z). Part of this work was supported by the Bill & Melinda Gates Foundation (BMGF-Calestous Juma Science Leadership Fellowship INV-036643) to YB. Part of the work was undertaken at UCLH/UCL who received a proportion of funding from the National Institute for Health Research (NIHR) University College London Hospitals Department of Health’s NIHR Biomedical Research Centre (BRC). ECW is supported by the Centre’s funding scheme. This work was supported jointly by the BRC and core funding from the Francis Crick Institute, which receives its funding from Cancer Research UK, the UK Medical Research Council, and the Wellcome Trust. EJC is supported by an MRC clinician scientist fellowship. DLVB is additionally supported by the Genotype-to-Phenotype National Virology Consortium (G2P-UK), Genotype-to-Phenotype 2 (G2P2-UK) and via UK Research and Innovation and the UK Medical Research Council.

This work was supported by the National Institute for Health Research University College London Hospitals Department of Health’s NIHR Biomedical Research Centre (BRC), as well as by the UK Research and Innovation and the UK Medical Research Council (MR/Y004337/1 to EW, MYW; MR/W005611/1 and MR/Y004205/1 to DVLB; and MR/X006751/1 to EJC), and by the Francis Crick Institute which receives its core funding from Cancer Research UK (CC2166, CC1283, CC1114, CC2230, CC2060, CC2041), the UK Medical Research Council (CC2166, CC1283, CC1114, CC2230, CC2060, CC2041), and the Wellcome Trust (CC2166, CC1283, CC1114, CC2230, CC2060, CC2041). This research was funded in part, by the Wellcome Trust [CC2166, CC1283, CC1114, CC2230, CC2060, CC2041 & 226142/Z/22/Z]. The funders of the study had no role in study design, data collection, data analysis, data interpretation, or writing of the report.

## Supplementary Figures & Tables

**Supplemental Figure 1.**
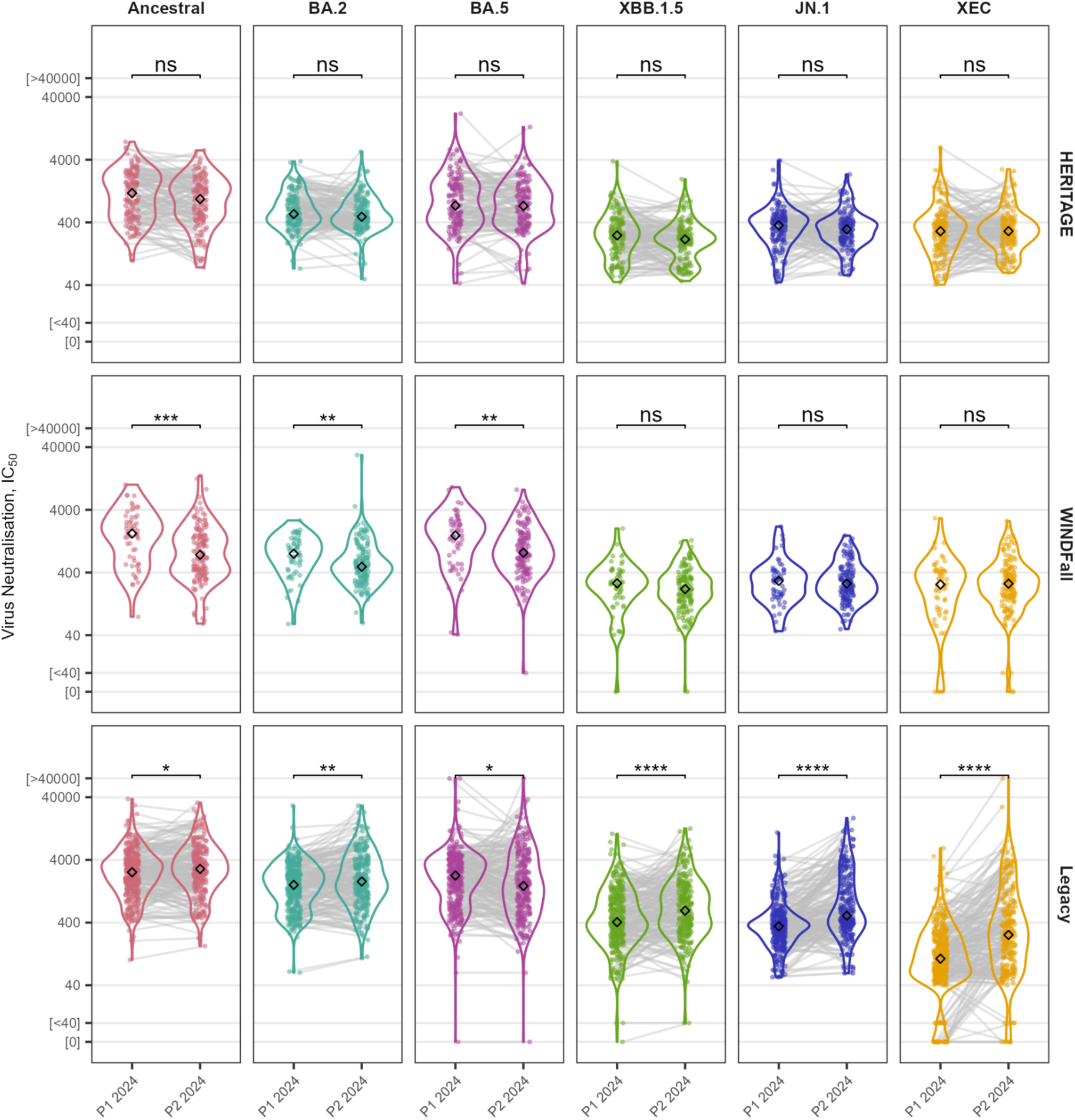
Within study variation in SARS-CoV-2 serological immunity during 2024. Serum neutralisation titres against SARS-CoV-2 variants (facet columns) from HERITAGE, WINDFall, and Legacy participants (facet rows) grouped into two timepoints in 2024: Period 1, April 1^st^ – August 18^th^; and Period 2, August 19^th^ – December 31^st^. Titres are plotted as Log_2_-transformed 50% inhibitory concentrations (IC_50_), the reciprocal of the serum dilution at which 50% of viral infection is inhibited. Grey lines connect paired sera from participants sampled at both time points (number of paired sera = 132 (HERITAGE), 0 (WINDFall), 190 (Legacy)). Significance was determined by two-tailed Wilcoxon rank-sum tests and is indicated by asterisks: * p < 0.05; ** p < 0.01; *** p < 0.001; **** P < 0.0001; ns, not significant (p ≥ 0.05).

**Supplemental Figure 2.**
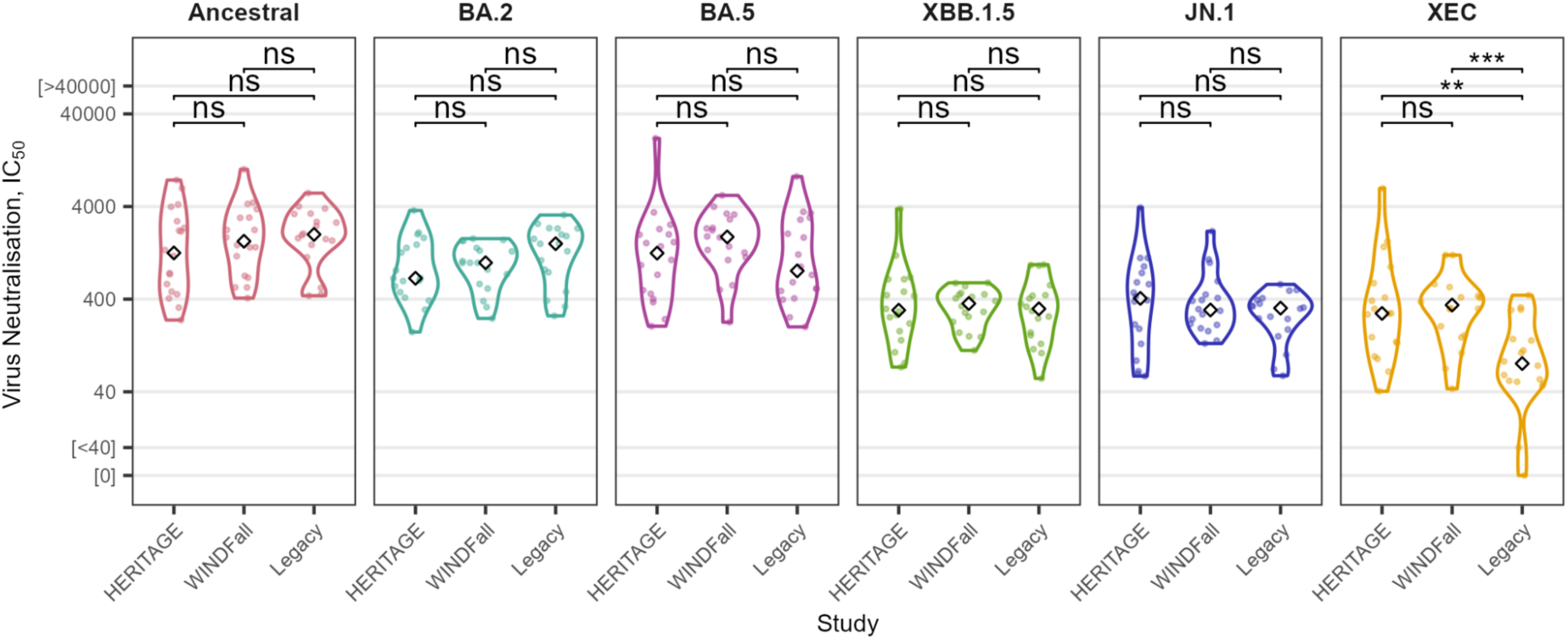
Spatial differences in antibody immunity in a matched sub-cohort during Period 1 2024. Serum neutralisation titres in Period 1 2024 (April 1^st^ – August 18^th^) from HERITAGE, WINDFall, and Legacy participants matched based on age group (≤35, 35-50, ≥50), sex, and number of vaccinations (≥2 doses & < 4 doses). Titres are plotted as Log_2_-transformed 50% inhibitory concentrations (IC_50_), the reciprocal of the serum dilution at which 50% of viral infection is inhibited. Significance was determined by two-tailed Wilcoxon rank-sum tests and is indicated by asterisks: * p < 0.05; ** p < 0.01; *** p < 0.001; **** P < 0.0001; ns, not significant (p ≥ 0.05).

**Supplemental Figure 3.**
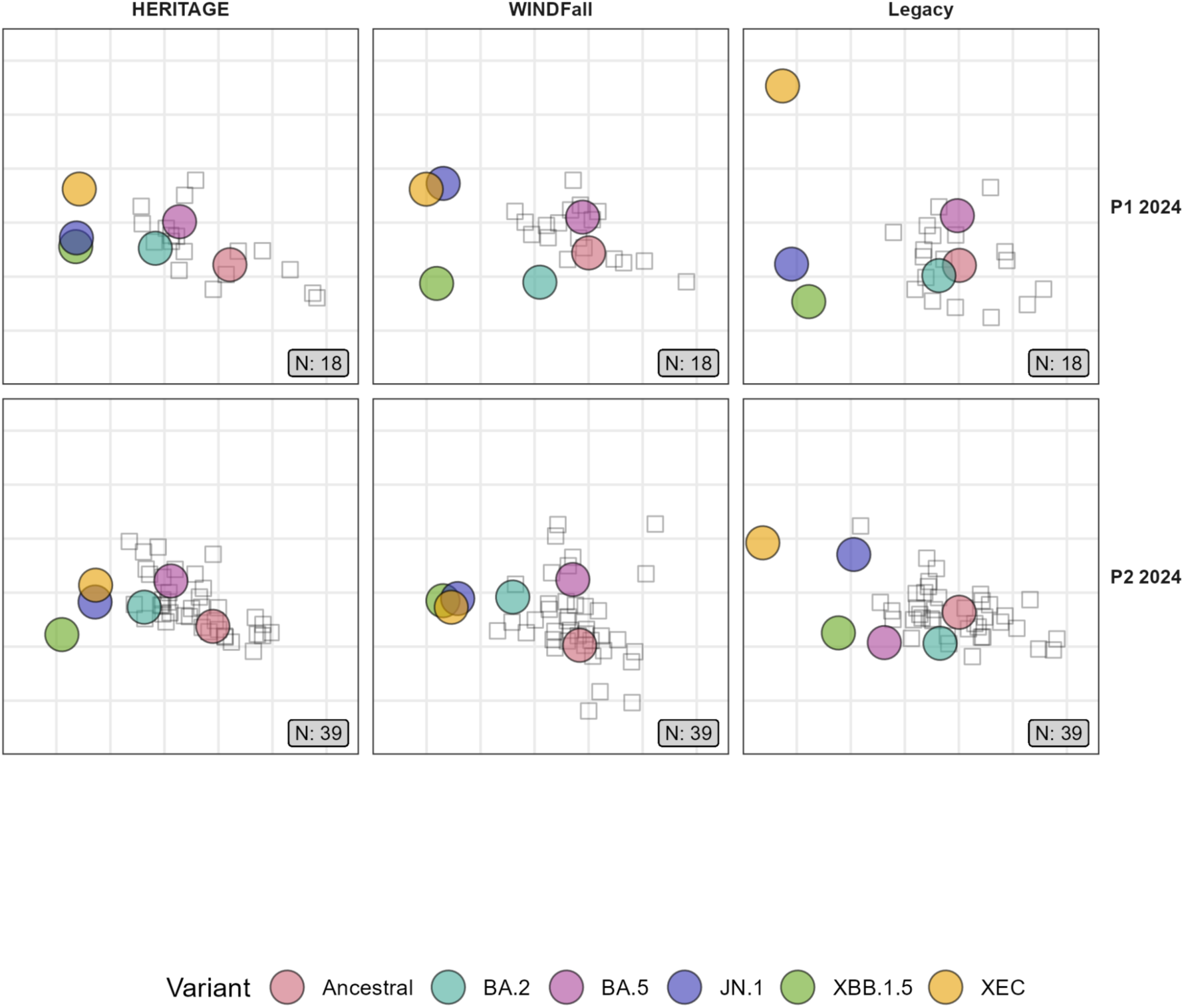
Geographic differences in antigenic space for matched sub-cohorts. Antigenic maps of SARS-CoV-2 variants for each study and timepoint in 2024 using matched sub-cohorts, with HERITAGE, WINDFall, and Legacy (columns left to right) and rows Period 1 (top) and Period 2 (bottom). See Table 4 & **Supplemental Table 1** for demographic information for the matched Period 2 and Period 1 sub-cohorts, respectively. Coloured circles denote virus antigens; black squares indicate individual sera; the N labels show the count of sera. Maps were generated in Racmacs; one grid unit represents a two-fold change in neutralization titre. Proximity between a serum and a variant reflects stronger neutralisation of that variant.

**Supplemental Table 1.** Participant demographics for Period 1 (P1, April 1st – August 18th 2024) when matched on age group (≤35, 35-50, ≥50), sex, and the number of SARS-CoV-2 vaccinations (≥2 doses & < 4 doses)

|  | HERITAGE, N = 18<sup>1</sup> | WINDFall, N = 18<sup>1</sup> | Legacy, N = 18<sup>1</sup> |
| --- | --- | --- | --- |
| Age | 40 [30-50] | 42 [31-55] | 45 [33-52] |
| Sex (Female) | 13 (72%) | 13 (72%) | 13 (72%) |
| BMI | NA [NA-NA] | 26.6 [22.1-32.2] | 22.3 [21.3-24.2] |
| Unknown | 18 | 2 | 0 |
| Vaccinations at sampling (N) |  |  |  |
| Median [25%-75%] | 2.00 [2.00-2.00] | 2.00 [2.00-2.75] | 3.00 [3.00-3.00] |
| Minimum-Maximum | 2.00-3.00 | 2.00-3.00 | 2.00-3.00 |
| Roche N result (Positive) | 17 (94%) | 13 (93%) | 18 (100%) |
| Unknown | 0 | 4 | 0 |
| First dose |  |  |  |
| AZD1222 | 9 (50%) | 14 (78%) | 5 (28%) |
| BNT162b2 | 9 (50%) | 4 (22%) | 13 (72%) |
| Second dose |  |  |  |
| AZD1222 | 9 (50%) | 14 (78%) | 5 (28%) |
| BNT162b2 | 9 (50%) | 4 (22%) | 13 (72%) |
| Third dose |  |  |  |
| AZD1222 | 2 (67%) | 0 (0%) | 0 (0%) |
| BNT162b2 | 1 (33%) | 5 (100%) | 14 (88%) |
| mRNA1273 | 0 (0%) | 0 (0%) | 1 (6.3%) |
| mRNA1273.214 | 0 (0%) | 0 (0%) | 1 (6.3%) |
| Unknown | 15 | 13 | 2 |
| Ethnicity |  |  |  |
| Asian | 0 (NA%) | 0 (0%) | 2 (11%) |
| Black | 0 (NA%) | 13 (81%) | 0 (0%) |
| Mixed | 0 (NA%) | 3 (19%) | 0 (0%) |
| Other | 0 (NA%) | 0 (0%) | 1 (5.6%) |
| White | 0 (NA%) | 0 (0%) | 15 (83%) |
| Unknown | 18 | 2 | 0 |

<sup>1</sup>Median [25%-75%]; n (%)

**Supplemental Table 2.**
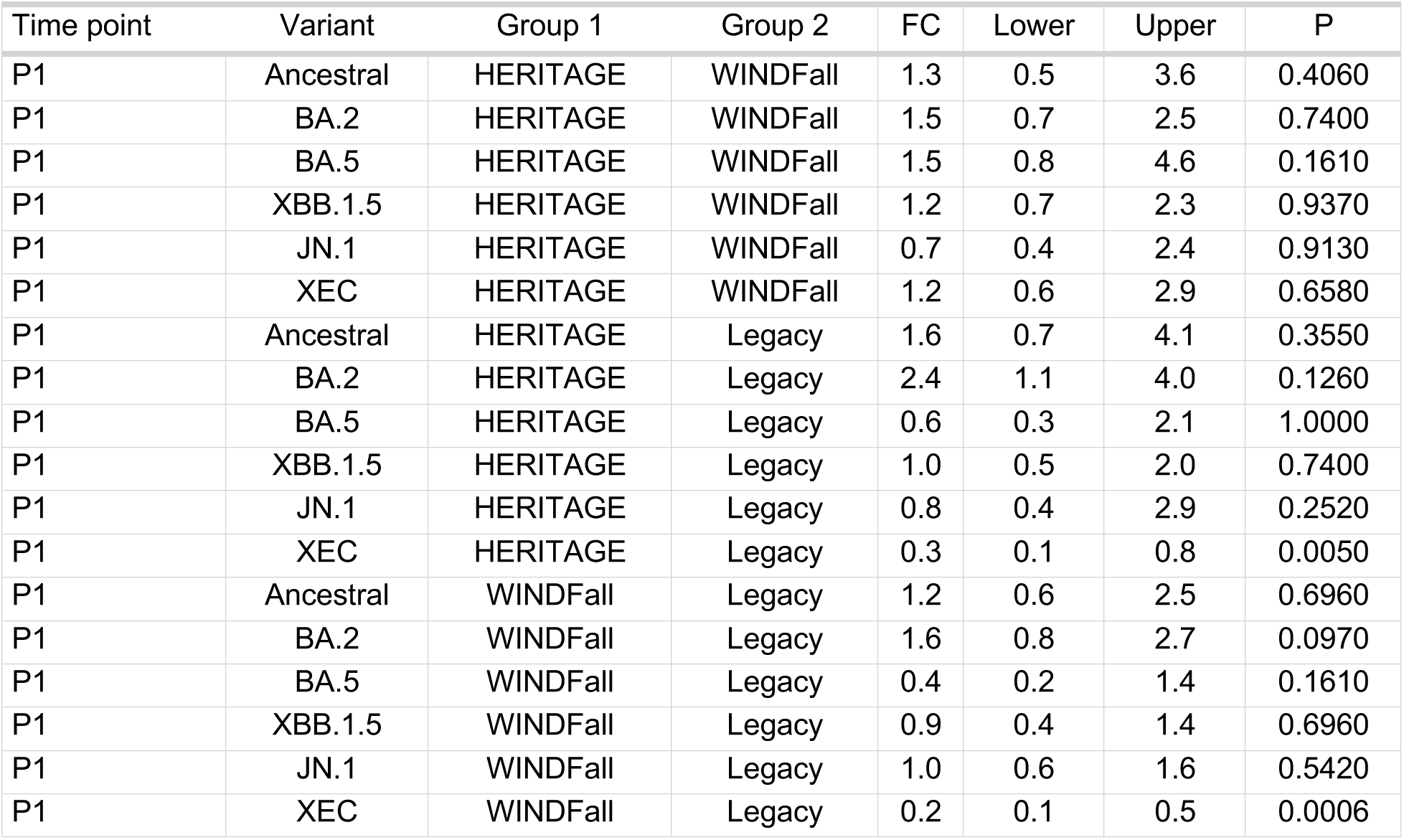
Fold change (FC) in serum neutralisation titres in a matched sub-cohort during Period 1 (P1, April 1st – August 18th 2024)

| Time point | Variant | Group 1 | Group 2 | FC | Lower | Upper | P |
| --- | --- | --- | --- | --- | --- | --- | --- |
| P1 | Ancestral | HERITAGE | WINDFall | 1.3 | 0.5 | 3.6 | 0.4060 |
| P1 | BA.2 | HERITAGE | WINDFall | 1.5 | 0.7 | 2.5 | 0.7400 |
| P1 | BA.5 | HERITAGE | WINDFall | 1.5 | 0.8 | 4.6 | 0.1610 |
| P1 | XBB.1.5 | HERITAGE | WINDFall | 1.2 | 0.7 | 2.3 | 0.9370 |
| P1 | JN.1 | HERITAGE | WINDFall | 0.7 | 0.4 | 2.4 | 0.9130 |
| P1 | XEC | HERITAGE | WINDFall | 1.2 | 0.6 | 2.9 | 0.6580 |
| P1 | Ancestral | HERITAGE | Legacy | 1.6 | 0.7 | 4.1 | 0.3550 |
| P1 | BA.2 | HERITAGE | Legacy | 2.4 | 1.1 | 4.0 | 0.1260 |
| P1 | BA.5 | HERITAGE | Legacy | 0.6 | 0.3 | 2.1 | 1.0000 |
| P1 | XBB.1.5 | HERITAGE | Legacy | 1.0 | 0.5 | 2.0 | 0.7400 |
| P1 | JN.1 | HERITAGE | Legacy | 0.8 | 0.4 | 2.9 | 0.2520 |
| P1 | XEC | HERITAGE | Legacy | 0.3 | 0.1 | 0.8 | 0.0050 |
| P1 | Ancestral | WINDFall | Legacy | 1.2 | 0.6 | 2.5 | 0.6960 |
| P1 | BA.2 | WINDFall | Legacy | 1.6 | 0.8 | 2.7 | 0.0970 |
| P1 | BA.5 | WINDFall | Legacy | 0.4 | 0.2 | 1.4 | 0.1610 |
| P1 | XBB.1.5 | WINDFall | Legacy | 0.9 | 0.4 | 1.4 | 0.6960 |
| P1 | JN.1 | WINDFall | Legacy | 1.0 | 0.6 | 1.6 | 0.5420 |
| P1 | XEC | WINDFall | Legacy | 0.2 | 0.1 | 0.5 | 0.0006 |

**Supplemental Table 3.** Fold change (FC) in serum neutralisation titres of HCoV-0C43 and HCoV-229E during either period of 2024.

| Time point | Variant | Group 1 | Group 2 | FC | Lower | Upper | P |
| --- | --- | --- | --- | --- | --- | --- | --- |
| P1 | OC43 | HERITAGE | WINDFall | 0.8 | 0.7 | 0.9 | 0.0020 |
| P1 | 229E | HERITAGE | WINDFall | 0.8 | 0.7 | 1.0 | 0.0050 |
| P1 | OC43 | HERITAGE | Legacy | 0.9 | 0.8 | 1.0 | 0.0010 |
| P1 | 229E | HERITAGE | Legacy | 0.8 | 0.7 | 0.9 | 0.0000 |
| P1 | OC43 | WINDFall | Legacy | 1.1 | 1.0 | 1.3 | 0.1980 |
| P1 | 229E | WINDFall | Legacy | 1.1 | 0.9 | 1.2 | 0.6610 |
| P2 | OC43 | HERITAGE | WINDFall | 0.6 | 0.6 | 0.7 | 0.0000 |
| P2 | 229E | HERITAGE | WINDFall | 0.6 | 0.5 | 0.6 | 0.0000 |
| P2 | OC43 | HERITAGE | Legacy | 1.0 | 0.9 | 1.1 | 0.8100 |
| P2 | 229E | HERITAGE | Legacy | 0.7 | 0.7 | 0.8 | 0.0000 |
| P2 | OC43 | WINDFall | Legacy | 1.6 | 1.4 | 1.7 | 0.0000 |
| P2 | 229E | WINDFall | Legacy | 1.3 | 1.2 | 1.5 | 0.0000 |

## Consortium membership

### Crick Serology Consortium

James Bazire, Guilia Dowgier, Agnieszka Hobbs, Phoebe Stevenson-Leggett, Rebecca Penn, Ruth Harvey, Murad Miah, Philip Bawumia, Mauro Miranda, Odiesia Daley, Nicola O’Reilly, Nicholas Wilson, Chris Cheshire, Karen Ambrose, Amy Strange, Gavin Kelly, Svend Kjaer

### the HERITAGE study team

Yakubu Alhassan, Vera M Kotey, Seth Agyemang, Adelaide K Sromani, Stephanie Darko, Randy Tackie, Aisha M Mohammed, Patricia Kaba, Ruth Kiome, David Hutchful, Lily Paemka, Emmanuella Amoako, Joyce M Ngoi, Aida Manu, Abdul Razak Quao, Akosua Ayisi, Kwame Amponsa-Achiano, Franklin Asiedu Bekoe, Esmy Kotey, Susan Amoako, Stephen Larbi Darkoh, Seyram Atukpa, Apetsi Ampiah, Silas Lawer, Charles Ansong, Frank Danquah, Wisdom Aveey

### Legacy Investigators

#### Early career researchers

Marianne Shawe-Taylor, Joshua Gahir, Hermaleigh Townsley, Suraiya Sharmin

#### Senior researchers

Vincenzo Libri, Padmasayee Papineni, Tumena Corrah, Richard Gilson, Rupert Beale, Nicola S Lewis, Sonia Gandhi, Charles Swanton, Steve Gamblin, Bryan Williams

### WINDFall Study Team

Jenene Cameron, Keisha Francis, Joyce Goode-Jones, Lydia Myles-Saunders, Yakima Philips-Crosdale, Renata Prospere, Samantha Shippey, Tamara Thompson

### WWW Consortium Team

Christèle Nguepou Tchopba, Irene Amoakoh Owusu, Michael Yakass; Sylvia S Opoku; Nikita Sahadeo, Anushka Ramjag, Arianne Brown-Jordan, Soren Nicholls, Vernie Ramkissoon; Gift Ahimbisibwe, Karen Ambrose, Emily Till, Max Schouten; Harriet Mears; Timothy W Russell

